# Zinc Supplementation in the management of acute diarrhea in high-income countries – A systematic review and meta-analysis

**DOI:** 10.1101/2024.07.09.24310071

**Authors:** Túlio Revoredo, Lucas Victor Alves, David Romeiro Victor, João Guilherme Bezerra Alves

## Abstract

The World Health Organisation (WHO) and the United Nations Children’s Fund (UNICEF) recommend zinc supplementation for children with diarrhea.

However, low- and middle-income countries (LMICs) conducted the majority of the studies supporting this recommendation. Although the mortality rate of acute diarrhoea in developed countries is low, diarrhoea leads to a high number of clinical care and hospital admissions, which represents a significant economic burden. This systematic review assessed the therapeutic benefits of zinc supplementation in the treatment of acute diarrhoea in children living in high-income countries. We conducted a literature search on the Medline, Embase, Cochrane, and Scielo databases to find published randomised controlled trials on zinc supplementation and acute diarrhoea in children residing in developed countries. We conducted a systematic literature search of the databases, uncovered 609 titles, and included 3 trials, totaling 620 treated children with acute diarrhoea, after reviewing abstracts and full manuscripts for inclusion and exclusion criteria. Two studies showed that zinc did not interfere with the duration of diarrhea. According to the Cochrane Risk of Bias RoB2, risk was considered low in two studies and some concerns in another. There was no statistically significant reduction in the mean RR for the occurrence of diarrheal episodes after 7 days of zinc supplement administration (0.4% vs. 0.6%; RR 0.73; 95% CI 0.28-1.92; p = 0.53; I2 = 16%). Zinc supplementation did not reduce the duration of acute diarrhoea among children living in developed countries.

**Whats is New:**

-Zinc supplementation did not reduce the duration of acute diarrhea among children living in developed countries.
-The anti-diarrheal effect of zinc is dependent on zinc deficiency.
-The WHO and UNICEF recommended regimen of therapeutic zinc should not include high-income countries.

**Whats is Known:**

-Zinc supplementation may reduce the duration and severity of diarrhea in poor countries
-The World Health Organization (WHO) and the United Nations International Children’s Emergency Fund (UNICEF) recommend zinc supplementation for children with acute diarrhea

## Introduction

Diarrhoea is the third leading cause of death in children 1–59 months of age, most of which occur in developing countries (1). According to the World Health Organization (WHO), acute diarrhoea is defined as the discharge of loose stools or liquid stools ≥ 3 times per day for ≥ 3 days and < 14 days (2). Diarrhoea can quickly lead to fluid and electrolyte loss and may be life-threatening, especially in young infants and malnourished children. Besides, diarrhoea promotes nutritional deficiencies, reduces immunity, and impairs growth and development (3, 4, 5).

Worldwide, zinc deficiency is common, but reports of severe deficiency are rare (6). Zinc is an important micronutrient for cellular growth, cellular differentiation, and metabolism (7). Zinc deficiency may limit immunity and impair resistance to infections. Many studies have shown that zinc supplementation may reduce the duration and severity of diarrhoea (8, 9, 10, 11). A systematic review of randomised controlled trials found that oral zinc supplementation significantly reduces the duration of diarrhoea; however, only one of the 18 included studies took place in a developed country (12). Another systematic review observed no consistent benefit with zinc trials to treat diarrhoea (13). A Cochrane systematic review suggests that zinc may be of benefit to children aged six months or more in areas where the prevalence of zinc deficiency or malnutrition is high (14).

The World Health Organisation (WHO) and the United Nations Children’s Fund (UNICEF) recommend zinc supplementation for children with acute diarrhoea (15), last updated on August 9, 2023 (https://www.who.int/tools/elena/interventions/zinc-diarrhoea#:~:text=WHO%20Recommendations,the%20age%20of%20six%20months). However, few studies were conducted in developed countries, thereby limiting the global WHO recommendations for diarrhea. This systematic review aims to assess the therapeutic benefits of zinc supplementation in the treatment of acute diarrhoea in children living in high-income countries.

## Methods

We developed this systematic review with meta-analysis in accordance with the Cochrane Handbook for Systematic Reviews of Interventions (16) and reported it using the updated Preferred Reporting Items for Systematic Reviews and Meta-Analyses (PRISMA) Checklist. We registered this study under Prospero (CRD42024516946). Ethical approval is not required for this study, as it is a systematic review.

We conducted a literature search using Medline, Embase, Cochrane, and Scielo electronic databases, without any language restrictions. We selected only randomised controlled trials (RCTs) conducted in developed countries. The search included trials that were published in any language. Two reviewers (TR and JGBA) assessed the eligibility of each record. Initially, we screened the title and abstract. At this stage, we excluded studies that were not RCTs, not conducted in developed countries, and did not include data on human subjects, acute diarrhoea, or oral zinc administration. We obtained complete articles to conduct further reviews of pertinent studies. A third reviewer (LVA) resolved any disagreements over the selection of studies.

We utilized the following keywords: “zinc”, “zinc supplementation”, “oral zinc”, “diarrhoea”, “acute diarrhoea”, “diarrhoea”, and “randomised controlled trial”, “randomised clinical trial”, or “randomised clinical trial”.

We used a form to extract relevant data from studies, including year, country, study design, population, setting, blinding, allocation concealment, sample size, intervention, and outcomes. Moreover, to conduct our statistical pooling, we solely extracted data from studies’ intention-to-treat analyses. Finally, we used the WebPlotDigitizer tool to extract pertinent data from Kaplan-Meier curves and incorporate it into our meta-analysis.

### Eligibility Criteria

According to the World Bank criteria, this review included RCTs conducted in developed countries, while it excluded studies conducted in low- and middle-income countries (LMIC). https://datahelpdesk.worldbank.org/knowledgebase/articles/906519-world-bank-country-and-lending-groups#:~:text=For%20the%20current%202024%20fiscal,those%20with%20a%20GNI%20per). We selected studies that only included children with acute diarrhoea. The intervention consists of oral zinc administration alone, without any combination. The primary outcome was the duration of diarrhea. Secondary outcomes were stool frequency (measured by the exact number of defecations recorded per day), vomiting duration, hospitalisation, and death from diarrhoea.

### Risk-of-Bias Assessment

The Risk-of-Bias RoB-2 Toll (17) evaluated the risk of bias of included RCTs based on seven domains: (1) the randomization process; (2) deviations from intended intervention; (3) missing outcome data; (4) outcome measurement; (5) selection of the reported results; (6) incomplete reporting; and (7) power calculation/sample size. Before assessment, the reviewers were trained. A third reviewer (LVA) resolved any disagreements.

### Statistical analysis

We compared dichotomous endpoints using risk ratios (RR) and 95% confidence intervals. P values ≥ 0.05 were considered significant for the rejection of the null hypothesis that there were no differences in effects between interventions. We adopted the Mantel-Haenszel test for dichotomous data. We used a DerSimonian and Laird random-effects model to incorporate the assumption that true effect sizes varied between studies.

Moreover, to measure and analyse heterogeneity, we utilised the Cochran Q test and I^2^ statistics. P values ≥ 0.10 were considered significant for the rejection of the null hypothesis that the studies shared a common true effect size. We used the I2 statistic to assess the percentage of variance in observed effect sizes due to heterogeneity. Due to the small number of included studies, we opted not to incorporate prediction intervals to assess heterogeneity or funnel plots to search for publication bias.

We used Cochrane’s Review Manager Web for statistical analysis.

## Results

Searching the following databases yielded a total of 629 studies: PubMed (305), Embase (214), Cochrane (101), and SciELO (9). After excluding duplicate manuscripts (303) and studies performed in LMIC countries (322), 4 studies showed potential relevance for the full analysis. We then screened the full texts of the remaining 4 articles for eligibility, excluding one because it did not perform the intervention with only zinc. As a result, this systematic review included three articles (18, 19, and 20) (Figure 1).

**Figure 1.**
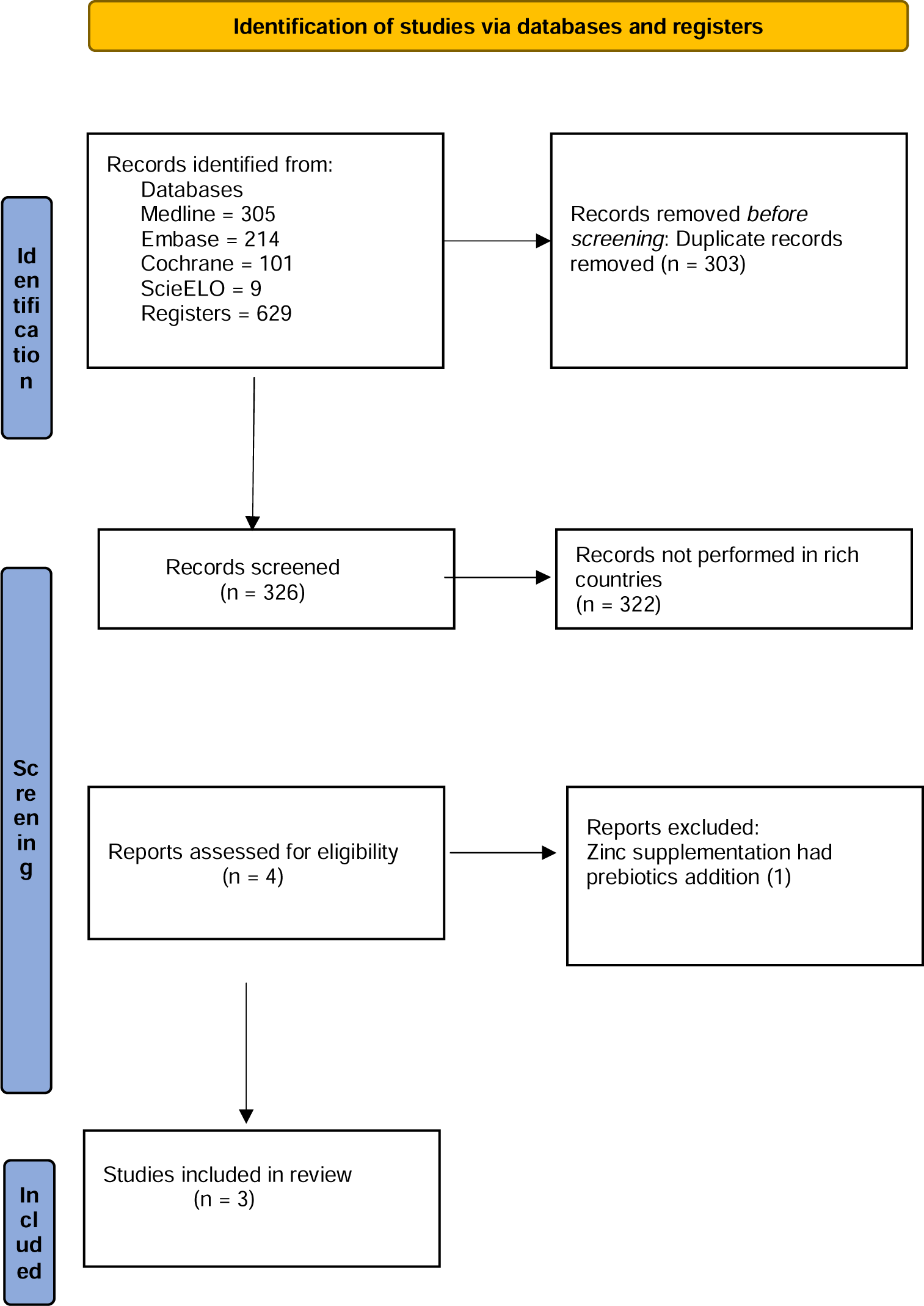
Flow chart.

The randomised controlled trial studies included 620 children with acute diarrhoea, with sample sizes ranging from 87 to 392. The age of participants ranged from 3 months to <11 years old. Table 1 displays the characteristics of the included studies.

**Table 1.** Characteristics of studies included in this systematic review.

| **Authors and year of publication** | **Country** | **Setting** | **Design** | **Allocation Concealment** | **Blinding** | **Sample size** | **Age (years)** | **Duration of diarrhea (p)** |
| --- | --- | --- | --- | --- | --- | --- | --- | --- |
| Valery et al. 2005 | Australia | Hospital | RCT | Yes | No | 392 | <11 years | 0.69 |
| Patro et al. 2010 | Poland | Hospital | RCT | Yes | Yes | 141 | 3 – 48 months | >0.05 |
| Crisinel et al. 2015 | Switzerland | Hospital | RCT | Yes | No | 87 | 3.1 - 50.3 months | 0.03 |

Legend: RCT – randomized controlled trial

Valery et al., in Australia, studied 392 Aboriginal children, < 11 years old, with diarrhoea supplemented with zinc, vitamin A, or combined zinc and vitamin A. They found no significant effect on the duration of diarrhea; the median diarrhoea duration after starting supplementation was 3.0 days for the Nd zinc supplemented and placebo groups (P values of 0.25 and 0.69, respectively).

Patro et al., in Poland, studied 69 children in the zinc-supplemented group compared with 72 children in the control group, and there was no significant difference in the duration of diarrhoea (P >.05). Similarly, they found no significant difference in secondary outcome measures (frequent stool and vomiting, intravenous fluid intake, and the number of children with diarrhea lasting >7 days).

Crisinel et al., in Switzerland, studied 87 children (median age 14 months; range 3.1–58.3); 42 received zinc supplementation and 45 received placebo. There was no difference in the duration or frequency of diarrhoea, but only 5% of the zinc group still had diarrhoea at 120 h of treatment, compared to 20% in the placebo group (P = 0.05). The average length of diarrhoea in zinc-treated patients was 47.5 hours (18.3–72 hours), which was significantly longer than the average length of diarrhoea in the placebo group (76.3 hours; IQR 52.8–137 hours) (P = 0.03). The frequency of diarrhoea was also lower in the zinc group (P = 0.02). The primary outcome was determined by intention-to-treat analysis, whereas the significant difference in median diarrhea duration was determined by per-protocol analysis.

We have assessed the risk of bias in the included studies. Table 2 displays the risk of bias in the included studies. We classified two studies as “low risk” (17, 18), and one as “moderate risk” due to missing data.

**Table 2.**
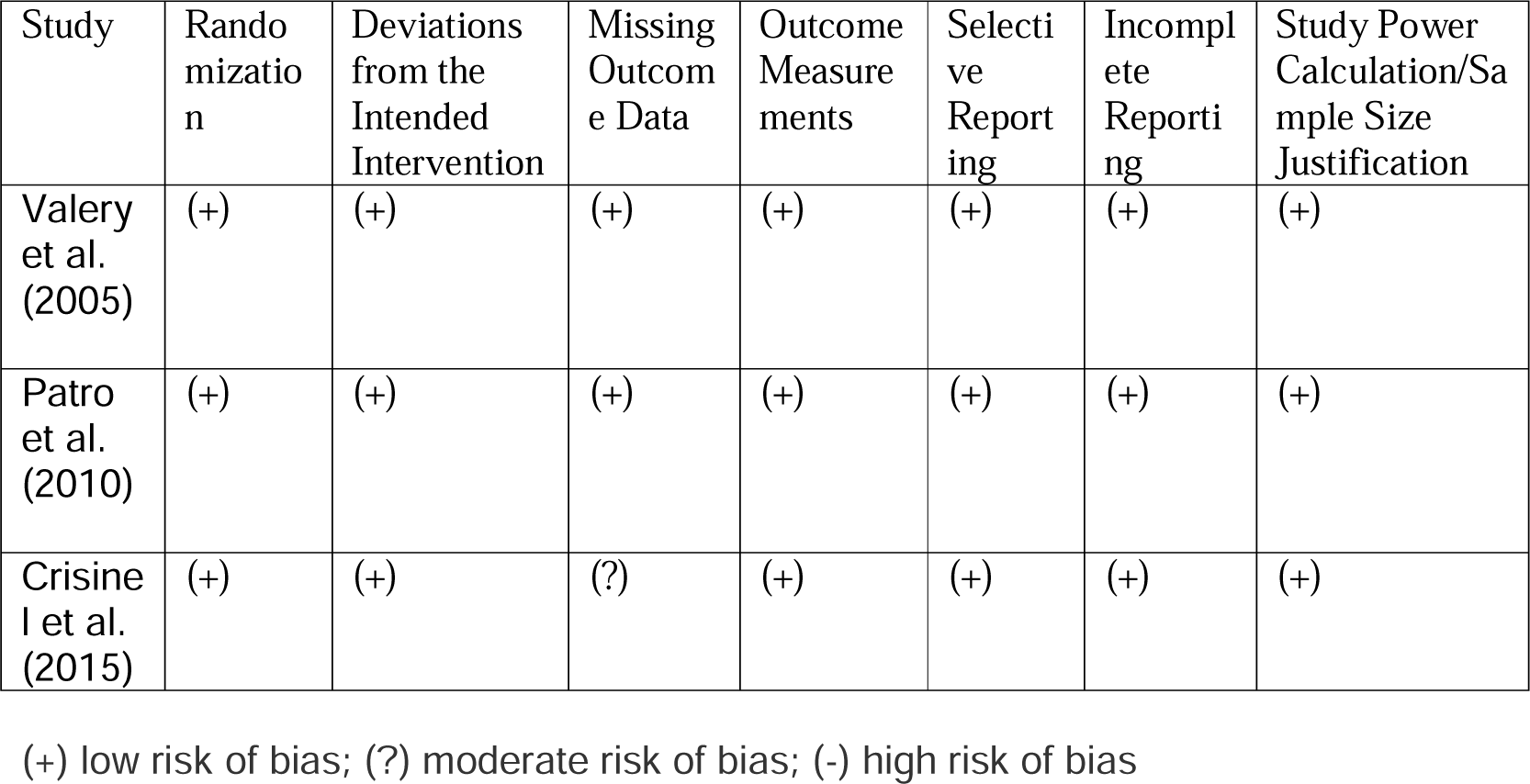
Assessment of the risk of bias based on the Cochrane Risk of Bias 2 checklist.

| Study | Rando<br>mizatio<br>n | Deviations<br>from the<br>Intended<br>Intervention | Missing<br>Outcom<br>e Data | Outcome<br>Measure<br>ments | Selecti<br>ve<br>Report<br>ing | Incompl<br>ete<br>Reporti<br>ng | Study Power<br>Calculation/Sa<br>mple Size<br>Justification |
| --- | --- | --- | --- | --- | --- | --- | --- |
| Valery<br>et al.<br>(2005) | (+) | (+) | (+) | (+) | (+) | (+) | (+) |
| Patro<br>et al.<br>(2010) | (+) | (+) | (+) | (+) | (+) | (+) | (+) |
| Crisine<br>l et al.<br>(2015) | (+) | (+) | (?) | (+) | (+) | (+) | (+) |

(+) low risk of bias; (?) moderate risk of bias; (-) high risk of bias

There was no statistically significant reduction in the mean RR in the occurrence of diarrheal episodes after 7 days of zinc supplement administration (0.4% vs. 0.6%; RR 0.73; 95% CI 0.28-1.92; p = 0.53; I2 = 16%; Figure 2). The mean RR for comparable studies could fall anywhere between 0.28 and 1.98, as represented by the 95% confidence interval. In regards to heterogeneity, we still can’t reject the null hypothesis that all studies share a common true effect size (p = 0.30) through the use of Cochrane’s Q statistic.

**Figure 2.**
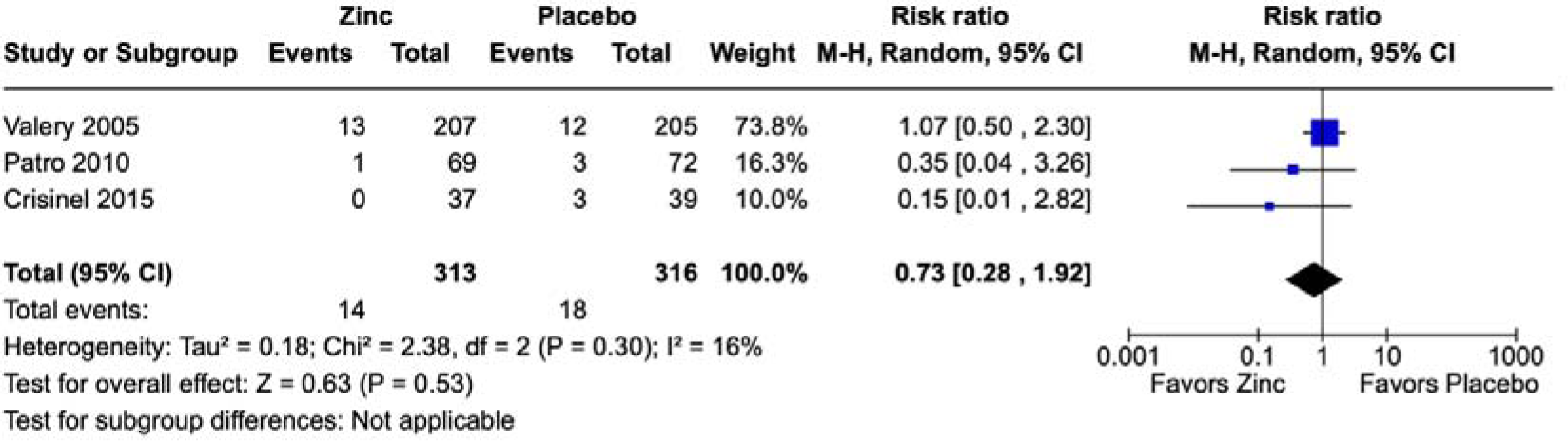
Forest plot for the occurrence of diarrheal episodes after 7 days of zinc supplementation.

## Discussion

The findings of this systematic review did not suggest the benefits of therapeutic zinc supplementation for diarrhoea among children in high-income countries, despite the detection of only three randomised controlled trials. In rich countries, the effects of zinc treatment, which include reductions in episode duration, stool output, stool frequency, and length of hospitalisation, were not the same as in studies performed in low- and middle-income countries. These results suggest that zinc therapy for diarrhoea seems not to be beneficial in high-income countries.

Patro et al. (18) observed no beneficial effect of zinc supplementation on diarrhoea duration or severity. They emphasise that they studied well-nourished and healthy children, who were therefore unlikely to be zinc deficient. T The authors attribute their inconsistent results with the majority of previously conducted trials, systematic reviews, and meta-analyses that have demonstrated an anti-diarrheal effect of zinc in children to the fact that these studies took place in countries with a medium or low Human Development Index (HDI), where malnutrition and zinc deficiency are significant issues. −10 They also stated that countries with a high or very high HDI have not conducted any studies evaluating the effects of zinc for the treatment of acute diarrhoea.

Valery et al. (19) came to the conclusion that hospitalised Aboriginal children in Australia may not benefit from zinc supplementation in the management of acute diarrhoea. However, they clarify that their findings may not apply to children suffering from malnutrition. They reported that their results for zinc supplementation differ from those in other settings because supplementation is only effective in populations with a low baseline level of these micronutrients. They add that their results suggest that zinc supplementation has a positive effect on stunted children.

This systematic review (20) included only one study that found a difference in the duration of diarrhoea: 47.5 h (18.3–72) in the zinc group and 76.3 h (52.8–137) in the placebo group (P = 0.03). However, this study has some limitations. First, they were unable to recruit the expected number of patients based on their calculations. Second, a large number of children were lost to follow-up; 60 patients, out of a total of 148 patients recruited for this study, were lost to follow-up without any available post-baseline data. Third, they had poor compliance, probably due to zinc’s metallic taste.

Another study (21), which was not part of our systematic review, showed that zinc supplementation had a positive effect on acute diarrhoea in a population of Italian children aged 3 to 36 months (for a duration less than 24 h). However, they used an oral rehydration solution (ORS) containing zinc and probiotics in addition to oral zinc for the intervention. For this reason, the question remains whether the effect was due to zinc or probiotics.

The possible zinc mechanisms of action against diarrhoea are not well understood. It may include improved absorption of water and electrolytes by the intestine, better regeneration of the gut epithelium, an increased number of enterocyte brush border enzymes, and an enhanced immune response (22, 23). However, it is questionable whether these actions are independent of zinc deficiency in the host. This systematic review pointed to this.

Many randomised controlled trials performed in LMIC countries have reported that oral zinc supplementation is effective in reducing the duration of acute diarrhoea (24, 25, 26, 27, 28, 29, 30), although some point to divergent results (31, 32). Based on these results, the World Health Organisation (WHO) and the United Nations Children’s Fund (UNICEF) recommend oral zinc supplementation as a universal treatment for all children with acute diarrhoea (WHO/UNICEF) (15). On the other side, according to the recommendations of the European Society for Paediatric Gastroenterology, Hepatology, and Nutrition/European Society for Paediatric Infectious Diseases (ESPGHAN/ESPID), there is not enough evidence to support its routine use in children with acute diarrhoea living in Europe, where zinc deficiency is rare (33). Although the mortality rate of acute diarrhoea in developed countries is low, diarrhoea leads to a high number of clinical care and hospital admissions, which represents a significant economic burden.

### Strengths and limitations

The strength of this study is its pioneering use of synthesised data from randomised clinical trials to evaluate the efficacy of zinc supplementation for the management of acute diarrhoea in children living in developed countries. This study also followed all the recommendations of the Cochrane Handbook for Systematic Reviews of Interventions (16) and reported according to the updated Preferred Reporting Items for Systematic Reviews and Meta-Analyses (PRISMA) Checklist.

In regards to our meta-analysis, one limitation stems from variations in the defined duration of diarrhoea episodes. While Patro et al. provided data solely for the prevalence of episodes lasting more than 7 days, (19) Valery et al. and Crisinel et al. reported data for episodes lasting 7 days or longer. (18, 20) Despite this inconsistency, we chose to combine this data for our pooling.

Additional limitations arise from variations in the age inclusion criteria across studies. Patro et al. included children up to a maximum of 4 years old (19), whereas Valery et al. extended their inclusion criteria to children up to 11 years old. (18) Thus, even if we couldn’t reject the null hypothesis that heterogeneity wasn’t present, this broader range likely does introduce heterogeneity into our meta-analysis. Furthermore, another potential source of heterogeneity arises from the study by Valery et al., as they included data on zinc supplementation among patients also receiving vitamin A supplements, not providing stratified data for patients receiving solely zinc supplementation against patients receiving solely the placebo. (18)

Finally, the primary constraint in our meta-analysis is undoubtedly the limited number of studies included. In random-effects meta-analysis, this poses a significant concern as it inhibits the accurate calculation of between-study variance. (34) Consequently, this may lead to unreliable estimates for the summary effect, its associated confidence interval, and the metrics pertaining to heterogeneity. (34) Hence, readers should exercise caution when interpreting our results and consider their limited scope. Nevertheless, despite these acknowledged limitations, conducting a meta-analysis is preferable to relying on an ad-hoc summary of the evidence. (34)

### Conclusions

Zinc supplementation did not reduce the duration of acute diarrhoea among children living in developed countries. This result supports the hypothesis that the anti-diarrheal effect of zinc is dependent on zinc deficiency. The WHO and UNICEF-recommended regimen of therapeutic zinc should include only low- and middle-income countries.

## Data Availability

All data produced are available online

## References

1 IHME. Global Burden of Diseases [Internet]. 2020 [cited 2021 Apr 14]. Available from: http://ghdx.healthdata.org/gbd-results-tool.

2 WHO. Diarrhoeal disease. https://www.who.int/news-room/fact-sheets/detail/diarrhoeal-disease

3 Das JK, Bhutta ZA. Global challenges in acute diarrhea. Curr Opin Gastroenterol. 2016;32(1):18–23. doi: 10.1097/MOG.0000000000000236.

4 Fleckenstein JM, Matthew Kuhlmann F, Sheikh A. Gastroenterol Acute Bacterial Gastroenteritis. Clin North Am. 2021;50(2):283–304. doi: 10.1016/j.gtc.2021.02.002. Epub 2021 Apr 23.

5 Kisenge R, Dhingra U, Rees CA, Liu E, Dutta A, Saikat D, Dhingra P, Somji S, Sudfeld C, Simon J, Ashorn P, Sazawal S, Duggan CP, Manji K. Risk factors for moderate acute malnutrition among children with acute diarrhoea in India and Tanzania: a secondary analysis of data from a randomized trial.

6 Bailey RL, West KP Jr, Black RE. The epidemiology of global micronutrient deficiencies. Ann Nutr Metab. 2015;66 Suppl 2:22–33. doi: 10.1159/000371618.

7 Costa MI, Sarmento-Ribeiro AB, Gonçalves AC. Zinc: From Biological Functions to Therapeutic Potential. Int J Mol Sci. 2023;24(5):4822. doi: 10.3390/ijms24054822.

8 Gregorio GV, Dans LF, Cordero CP, Panelo CA. Zinc supplementation reduced cost and duration of acute diarrhea in children. J Clin Epidemiol. 2007;60(6):560–6. doi: 10.1016/j.jclinepi.2006.08.004.

9 Zou TT, Mou J, Zhan X. Zinc supplementation in acute diarrhea. Indian J Pediatr. 2015;82(5):415–20. doi: 10.1007/s12098-014-1504-6.

10 Lamberti LM, Walker CL, Chan KY, Jian WY, Black RE. Oral zinc supplementation for the treatment of acute diarrhea in children: a systematic review and meta-analysis. Nutrients. 2013;5(11):4715–40. doi: 10.3390/nu5114715.

11 Lazzerini M. Oral zinc provision in acute diarrhea. Curr Opin Clin Nutr Metab Care. 2016;19(3):239–43. doi: 10.1097/MCO.0000000000000276.

12 Galvao TF, Thees MFRS, Pontes RF, Silva MT, Pereira, MG. Zinc supplementation for treating diarrhea in children: a systematic review and meta-analysis. Rev Panam Salud Publica 2013;33(5):370–7. doi: 10.1590/s1020-49892013000500009.

13 Pavlinac PB, Brander RL, Atlas HE, John-Stewart GC, Denno DM, Walson JL. Interventions to reduce post-acute consequences of diarrheal disease in children: a systematic review. BMC Public Health. 2018;18(1):208. doi: 10.1186/s12889-018-5092-7.

14 Lazzerini M, Wanzira H. Oral zinc for treating diarrhoea in children. Cochrane Database Syst Rev. 2016;12(12):CD005436. doi: 10.1002/14651858.CD005436.pub5.

15 WHO/UNICEF Clinical management of acute diarrhoea. Geneva: WHO; 2004. Report No.: WHO/FCH/CAH 04.7.

16 Higgins JPT, Thomas J, Chandler J, et al, Eds. Cochrane handbook for systematic reviews of interventions. Version 6.3, 2022, Cochrane. Accessed 4 February 2024. Available from www.training.cochrane.org/handbook

17 Sterne J.A.C., Savović J., Page M.J., Elbers R.G., Blencowe N.S., Boutron I., Cates C.J., Cheng H.-Y., Corbett M.S., Eldridge S.M., et al. RoB 2: A revised tool for assessing risk of bias in randomised trials. BMJ. 2019;366:l4898. doi: 10.1136/bmj.l4898.

18 Valery PC, Torzillo PJ, Boyce NC, White AV, Stewart PA, Wheaton GR, Purdie DM, Wakerman J, Chang AB. Zinc and vitamin A supplementation in Australian Indigenous children with acute diarrhoea: a randomised controlled trial. Med J Aust. 2005;182(10):530–5. doi: 10.5694/j.1326-5377.2005.tb00021.x.

19 Patro B, Szymański H, Szajewska H. Oral zinc for the treatment of acute gastroenteritis in Polish children: a randomized, double-blind, placebo-controlled trial. J Pediatr. 2010;157(6):984–988.e1. doi: 10.1016/j.jpeds.2010.05.049.

20 Crisinel PA, Verga ME, Kouame KS, Pittet A, Rey-Bellet CG, Fontaine O, Di Paolo ER, Gehri M. Demonstration of the effectiveness of zinc in diarrhoea of children living in Switzerland. Eur J Pediatr. 2015;174(8):1061–7. doi: 10.1007/s00431-015-2512-x.

21 Passariello A, Terrin G, De Marco G, Cecere G, Ruotolo S, Marino A, Cosenza L, Tardi M, Nocerino R, Berni Canani R. Efficacy of a new hypotonic oral rehydration solution containing zinc and prebiotics in the treatment of childhood acute diarrhea: a randomized controlled trial. J Pediatr 2011;158:288–292, e.

22 Berni Canani R, Buccigrossi V, Passariello A Mechanisms of action of zinc in acute diarrhea. Curr Opin Gastroenterol. 2011;27(1):8-12. doi: 10.1097/MOG.0b013e32833fd48a.

23 Hassan A, Sada KK, Ketheeswaran S, Dubey AK, Bhat MS.Cureus Role of Zinc in Mucosal Health and Disease: A Review of Physiological, Biochemical, and Molecular Processes. 2020;12(5):e8197. doi: 10.7759/cureus.8197.

24 Roy SK, Tomkins AM, Akramuzzaman SM, Behrens RH, Haider R, Mahalanabis D, Fuchs G. Randomised controlled trial of zinc supplementation in malnourished Bangladeshi children with acute diarrhoea. Arch Dis Child. 1997;77(3):196–200. doi: 10.1136/adc.77.3.196.

25 Strand TA, Chandyo RK, Bahl R, Sharma PR, Adhikari RK, Bhandari N, Ulvik RJ, Mølbak K, Bhan MK, Sommerfelt H. Effectiveness and efficacy of zinc for the treatment of acute diarrhea in young children. Pediatrics. 2002;109(5):898–903. doi: 10.1542/peds.109.5.898.

26 Bhatnagar S, Bahl R, Sharma PK, Kumar GT, Saxena SK, Bhan MK. Zinc with oral rehydration therapy reduces stool output and duration of diarrhea in hospitalized children: a randomized controlled trial. J Pediatr Gastroenterol Nutr. 2004;38(1):34-40. doi: 10.1097/00005176-200401000-00010.

27 Trivedi SS, Chudasama RK, Patel N. Effect of Zinc Supplementation in Children with Acute Diarrhea: Randomized Double Blind Controlled Trial. Gastroenterology Res. 2009;2(3):168–174. doi: 10.4021/gr2009.06.1298.

28 Yazar AS, Güven Ş, Dinleyici EÇ. Effects of zinc or synbiotic on the duration of diarrhea in children with acute infectious diarrhea. Turk J Gastroenterol. 2016;27(6):537–540. doi: 10.5152/tjg.2016.16396.

29 Dhingra U, Kisenge R, Sudfeld CR, Dhingra P, Somji S, Dutta A, Bakari M, Deb S, Devi P, Liu E, Chauhan A, Kumar J, Semwal OP, Aboud S, Bahl R, Ashorn P, Simon J, Duggan CP, Sazawal S, Manji K. Lower-Dose Zinc for Childhood Diarrhea - A Randomized, Multicenter Trial. N Engl J Med. 2020;383(13):1231–1241. doi: 10.1056/NEJMoa1915905.

30 Rerksuppaphol L, Rerksuppaphol S. Efficacy of zinc supplementation in the management of acute diarrhoea: a randomised controlled trial. Paediatr Int Child Health. 2020;40(2):105–110. doi: 10.1080/20469047.2019.1673548.

31 Negi R, Dewan P, Shah D, Das S, Bhatnagar S, Gupta P. Oral zinc supplements are ineffective for treating acute dehydrating diarrhoea in 5-12-year-olds. Acta Paediatr. 2015;104(8):e367–71. doi: 10.1111/apa.12645.

32 Wadhwa N, Natchu UC, Sommerfelt H, Strand TA, Kapoor V, Saini S, Kainth US, Bhatnagar S. ORS containing zinc does not reduce duration or stool volume of acute diarrhea in hospitalized children. J Pediatr Gastroenterol Nutr. 2011;53(2):161–7. doi: 10.1097/MPG.0b013e318213ca55.

33 Guarino A, Albano F, Ashkenazi S, Gendrel D, Hoekstra JH, Shamir R, et al., Expert Working Group. The ESPGHAN/ESPID evidenced-based guidelines for the management of acute gastroenteritis in children in Europe. J Pediatr Gastroenterol Nutr 2008;46(Suppl 2):S81–122.

34 Hedges, L. V., Higgins, J. P. T., Rothstein, H. R., & Borenstein, M. (2011, August 24).Introduction to Meta-Analysis. John Wiley & Sons.

